# Prioritizing countries for TB vaccine readiness research using a global stakeholder-centric approach

**DOI:** 10.1101/2025.05.02.25326875

**Authors:** Michelle M. Gill, Rupali Limaye, Puck T. Pelzer, Mike Frick, Andrew D. Kerkhoff

## Abstract

**Background:** The promise of new tuberculosis (TB) vaccine candidates prompts the need for research on vaccine demand and health system readiness in diverse country settings with high TB burdens to help ensure effective and equitable vaccine deployment. Prior to vaccine introduction, we developed a structured approach to prioritizing countries for TB vaccine readiness research by combining stakeholder preferences, elicited through best-worst scaling (BWS) with an analytical hierarchy process (AHP) framework.

**Methods:** We conducted a self-administered electronic survey targeted to TB vaccine stakeholders involved in vaccine development, advocacy, and implementation across 23 of the 24 USAID TB priority countries. The survey included BWS to determine the relative importance of 17 criteria for country selection. Stakeholders were recruited using an existing email list, a ‘snowball’ approach, and recommendations from TB experts. In a series of 13 choice tasks, respondents selected what they felt were the most and least important criteria from four randomly generated criteria. The weights derived through BWS for each criterion were combined with country-specific scores for each criterion using publicly available data to determine the overall prioritization score for each country.

**Results:** Of 427 stakeholders, 115 (26%) completed the survey; 88% were from TB priority countries. Participants represented civil society/advocacy groups (34%), international nongovernmental organizations (21%), national TB Programs (13%), and academic/research institutions (17%). Sixteen of 17 criteria were identified as ‘important’ using BWS. Overall country TB burden (weight=11.1) and TB-related political will (weight=10.3) were the most important, followed by burden of TB-related deaths (weight=7.9), health systems strength (weight=7.5), and adult COVID-19 coverage (weight=7.4). The five countries with the highest prioritization scores were in sub-Saharan Africa. Three of them were selected alongside the highest-scoring country from South Asia, Europe and Central Asia, and East Asia as priority research settings in pursuit of regional diversity.

**Conclusion:** This study demonstrates the successful use of an inclusive, objective approach to be considered alongside other contextual factors for prioritizing countries for future TB vaccine readiness research. The AHP, especially when combined with BWS, is a practical and transparent approach that can be applied to support evidence-based funding decisions in the global public health field.

## INTRODUCTION

With several tuberculosis (TB) vaccine candidates for adults and adolescents now in phase 3 trials, the global fight against TB may soon enter a critical phase, as these vaccines could become available in the near future [1, 2]. This development, combined with lessons learned from vaccine introduction and delivery during the global COVID-19 pandemic, underscores the importance of strategic, coordinated planning for implementation readiness, to help ensure availability, accessibility, and acceptability of TB vaccines [3, 4]. Assessing vaccine demand and health system readiness factors in diverse countries with high TB burdens will be crucial for ensuring effective vaccine deployment and equitable impact when these vaccines become available.

Prior to new vaccine introduction, there is not yet a clear signal of demand for a new product and the vaccine pipeline and policy pathway may not be well understood, thus limiting available resources [5]. Therefore, this situation necessitates a careful and strategic process for selecting countries in which to undertake TB vaccine readiness research, considering the epidemiological burden of TB, health systems strength, political will, and community demand [4]. Such an approach aims to ensure that country-specific TB vaccine readiness research is targeted to maximize lessons learned within and across various settings. This will be particularly informative when developing TB vaccine delivery strategies for adults and adolescents given their lack of inclusion in most expanded programs on immunization (EPI) in high TB burden countries and their more limited interactions with the health system compared to infants [6].

The prioritization and allocation of global public health funding for implementation research have often been governed by methods that fall short of objectivity and transparency [7, 8]. Decisions can be influenced by top-down directives, such as political motives or historical funding patterns, which may not accurately reflect dynamic and context-specific public health needs [9]. This reliance on traditional prioritization and funding practices may exclude local experts and overshadow unbiased evaluations, creating a misalignment between available funding and the most pressing health challenges [9, 10]. The adoption of transparent, inclusive, and objective methods and processes for funding and prioritization is imperative for evidence-based decision-making and extends beyond TB vaccine research to broader global health initiatives.

Meaningful stakeholder engagement for TB vaccine development and implementation—including Government policymakers, affected persons and communities, civil society organizations, researchers, and healthcare workers—is pivotal to overcoming these issues and addressing complex global public health challenges [4, 11]. A stakeholder-centric approach not only can enhance transparency and equity in the funding prioritization and decision-making process but also ensures that strategies are closely aligned with the specific needs of diverse settings [8, 12]. Such an approach bridges the gap between high-level planning and ground-level realities, potentially leading to more effective and sustainable public health programs and activities.

Against this backdrop, we developed a structured and transparent approach to country prioritization for TB vaccine readiness research by integrating stakeholder preferences within a decision-making framework as part of the United States Agency for International Development (USAID)-funded Supporting, Mobilizing, and Accelerating Research for Tuberculosis Elimination (SMART4TB) project. We applied the Analytical Hierarchy Process (AHP) - a technique that provides a structured way to organize and assess complex decisions - that has been underutilized in global public health [13–15]. In conjunction with AHP, we integrated an online Best-Worst Scaling (BWS) exercise - a choice-based preference elicitation method - to enhance the intuitiveness and accessibility of indicating the relative importance of prioritization criteria to stakeholders. We describe the process used to design and implement this framework for selecting countries for TB vaccine readiness research [16–18].

## METHODS

### Design

This study used an AHP framework for country prioritization in future TB vaccine readiness research. Given the anticipated availability of new TB vaccines within 5-7 years, this prioritization process was needed to strategically plan and initiate implementation research that will support effective vaccine introduction [1–3]. A cross-sectional, self-administered survey was delivered through an electronic platform primarily to global, regional, and national stakeholders involved in TB vaccine development, advocacy, and implementation.

### AHP Overview and BWS Rationale

The overall goal of the AHP was to prioritize six countries among 24 USAID high TB burden countries to conduct multi-year, mixed-methods TB vaccine readiness research to maximize efficient use of funding. Our framework utilized a structured multi-step process, described in detail below: (1) defining the goal, (2) developing the hierarchy of the problem, including identifying the criteria, (3) determining criteria weights using a BWS among stakeholders, (4) determining country-specific scores from measurable, publicly available data and the BWS-derived weights, (5) synthesizing the results to determine country-specific overall prioritization scores, and (6) making final country selections (Figure 1).

**Figure 1.**
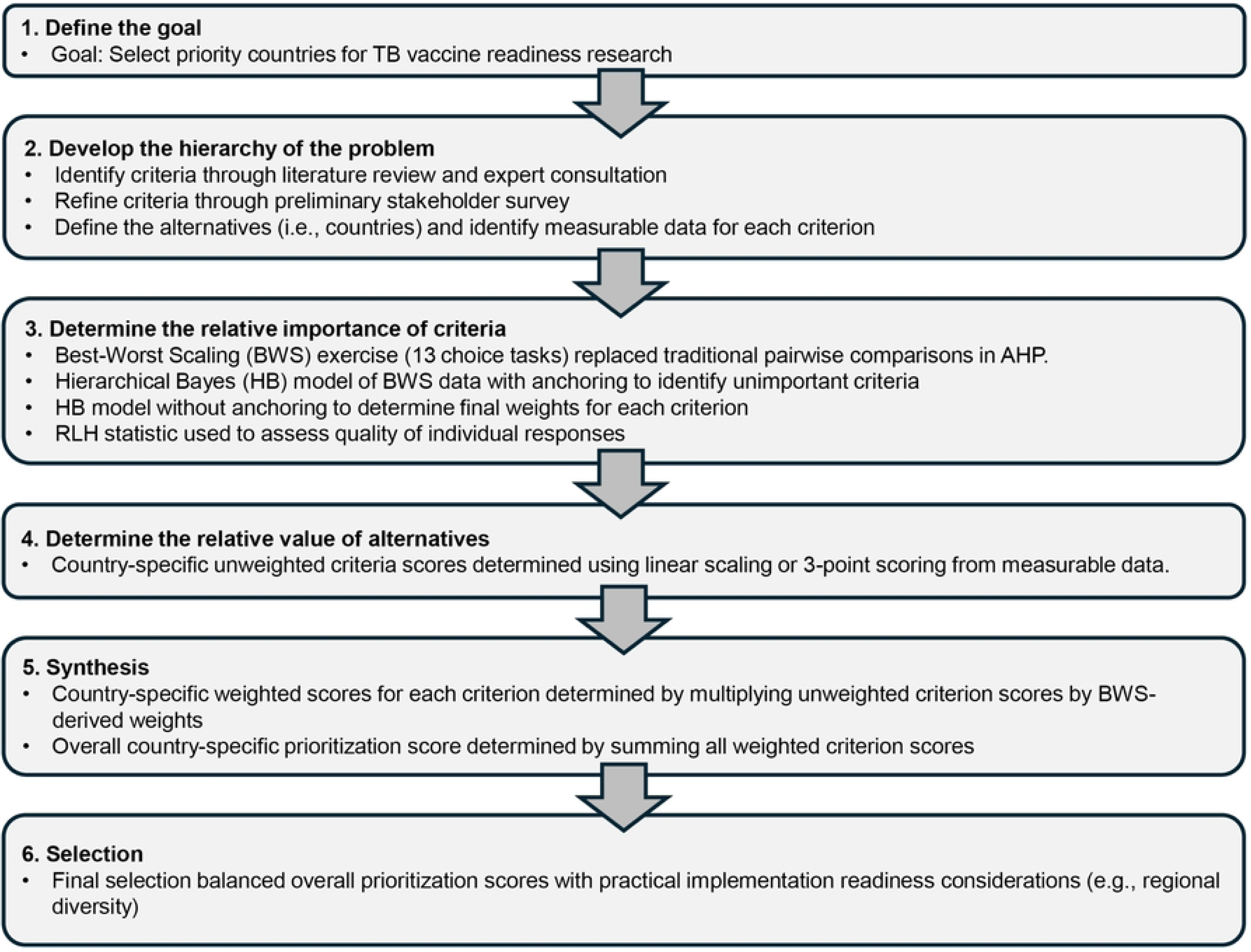
Adapted analytical hierarchy process framework incorporating best-worst scaling

To ensure the survey’s accessibility and reliability for participants who are non-native English speakers or have varying educational backgrounds, a type 1 BWS (object case) was chosen for criterion weight derivation. Unlike discrete choice experiments which are limited in the number of attributes that can typically be assessed (e.g., up to 7) and require attributes to have associated levels, a type 1 BWS was well-suited to derive and compare the relative importance of a large number of criteria [19]. This approach replaced the traditional AHP for determining criteria weights, which requires many grid-based pairwise comparisons, where respondents must score the importance of all criteria relative to all others using a scale from 1 (equal importance) to 9 (extreme importance), and reciprocally, from 1/9 to 1 for the inverse comparison [20, 21]. This can be time-consuming, cognitively demanding, especially when dealing with many criteria, and potentially confusing due to its wide relative importance scale range and reciprocal scoring system. In contrast, BWS simplifies this process by presenting several questions, each with a manageable set of 4-5 criteria, from which respondents are asked to identify the ‘best’ (most important) and ‘worst’ (least important) criteria. Thus, BWS could reduce cognitive load due to its straightforward ‘best’/‘worst’ selection for each question, potentially leading to more robust and reliable estimates [17, 18, 22].

### Identification of Criteria for Prioritization

To identify decision-making criteria, an initial list was informed by a review of the existing literature [23, 24] and expert consultations within the SMART4TB consortium via email and virtual meetings. To ensure the criteria to be assessed were comprehensive and identified through a collaborative process, in May 2023 a preliminary survey was sent to a small group of regional and global SMART4TB consortium members and external stakeholders involved in areas such as TB vaccine introduction, policy, and market analysis for their feedback (n=57).

Respondents (n=15, 26%) reviewed the proposed criteria and indicated whether they should be retained or removed. Respondents were also asked to indicate their familiarity with specific countries’ TB landscape and willingness to assist with refining target stakeholder lists for the main BWS survey.

The final list of 17 criteria assessed in the BWS survey, along with their respective data sources, are summarized in Table 1. Preliminary survey responses led to the removal of two criteria (adult Bacille Calmette-Guérin [BCG] vaccination and child/adolescent COVID-19 vaccination coverage) were recommended for removal by >50% of preliminary survey participants.

**Table 1.**
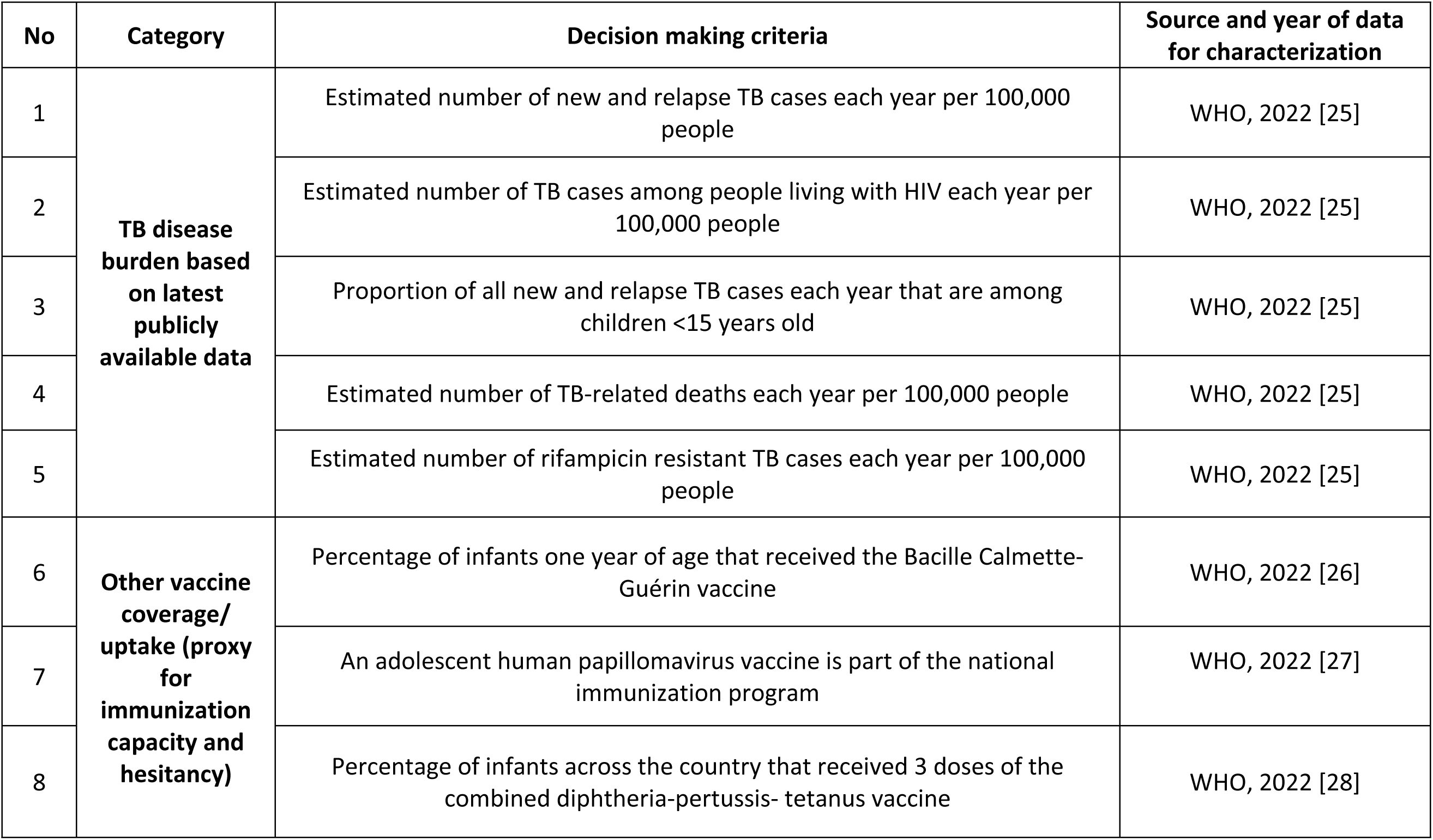
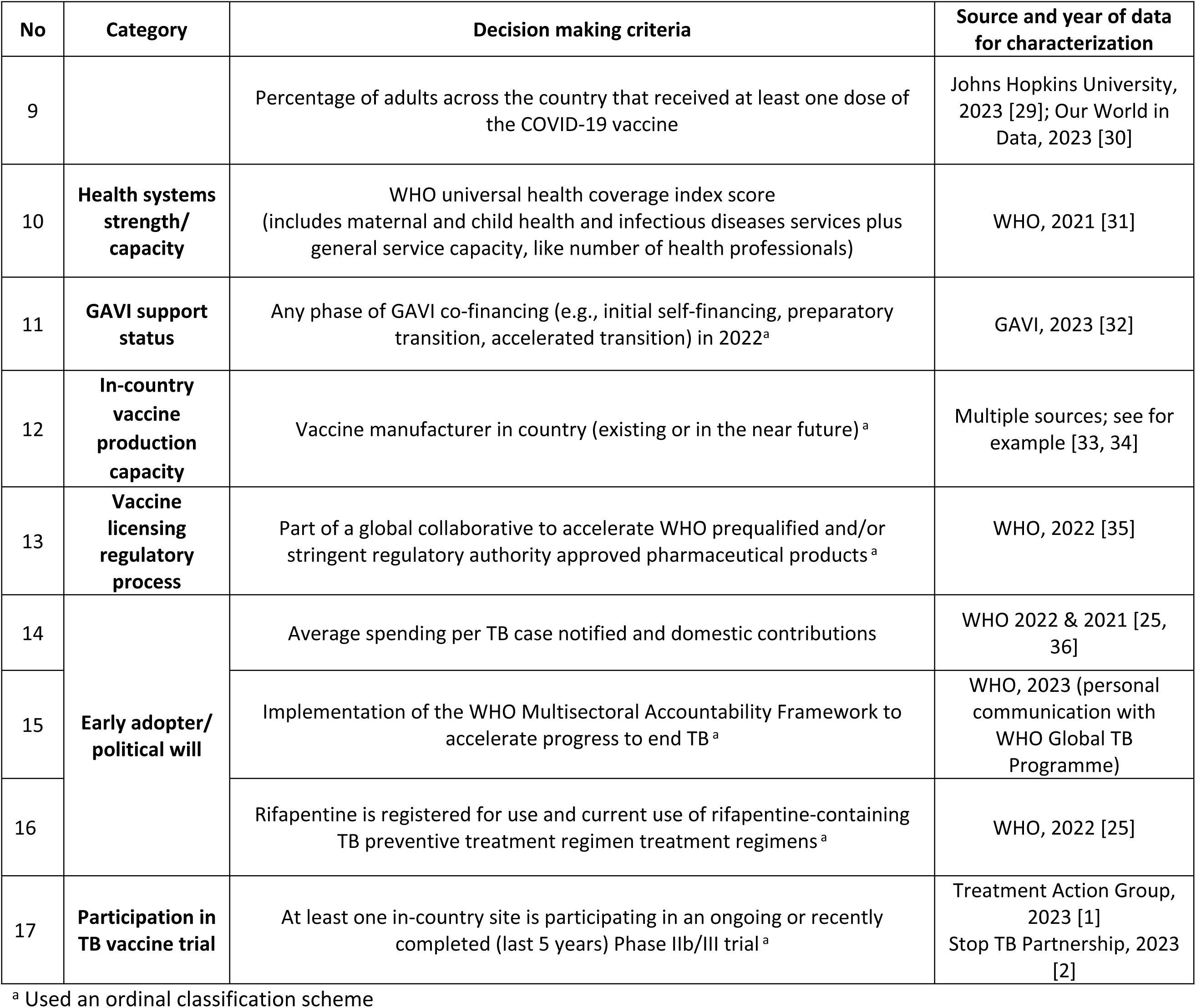
Best-worst scaling decision-making criteria used to determine country selection.

| No | Category | Decision making criteria | Source and year of data for characterization |
| --- | --- | --- | --- |
| 1 | **TB disease burden based on latest publicly available data** | Estimated number of new and relapse TB cases each year per 100,000 people | WHO, 2022 [25] |
| 2 |  | Estimated number of TB cases among people living with HIV each year per 100,000 people | WHO, 2022 [25] |
| 3 |  | Proportion of all new and relapse TB cases each year that are among children <15 years old | WHO, 2022 [25] |
| 4 |  | Estimated number of TB-related deaths each year per 100,000 people | WHO, 2022 [25] |
| 5 |  | Estimated number of rifampicin resistant TB cases each year per 100,000 people | WHO, 2022 [25] |
| 6 | **Other vaccine coverage/ uptake (proxy for immunization capacity and hesitancy)** | Percentage of infants one year of age that received the Bacille Calmette-Guérin vaccine | WHO, 2022 [26] |
| 7 |  | An adolescent human papillomavirus vaccine is part of the national immunization program | WHO, 2022 [27] |
| 8 |  | Percentage of infants across the country that received 3 doses of the combined diphtheria-pertussis- tetanus vaccine | WHO, 2022 [28] |
| 9 |  | Percentage of adults across the country that received at least one dose of the COVID-19 vaccine | Johns Hopkins University, 2023 [29]; Our World in Data, 2023 [30] |
| 10 | **Health systems strength/capacity** | WHO universal health coverage index score (includes maternal and child health and infectious diseases services plus general service capacity, like number of health professionals) | WHO, 2021 [31] |
| 11 | **GAVI support status** | Any phase of GAVI co-financing (e.g., initial self-financing, preparatory transition, accelerated transition) in 2022<sup>a</sup> | GAVI, 2023 [32] |
| 12 | **In-country vaccine production capacity** | Vaccine manufacturer in country (existing or in the near future)<sup>a</sup> | Multiple sources; see for example [33, 34] |
| 13 | **Vaccine licensing regulatory process** | Part of a global collaborative to accelerate WHO prequalified and/or stringent regulatory authority approved pharmaceutical products<sup>a</sup> | WHO, 2022 [35] |
| 14 | **Early adopter/political will** | Average spending per TB case notified and domestic contributions | WHO 2022 & 2021 [25, 36] |
| 15 |  | Implementation of the WHO Multisectoral Accountability Framework to accelerate progress to end TB<sup>a</sup> | WHO, 2023 (personal communication with WHO Global TB Programme) |
| 16 |  | Rifapentine is registered for use and current use of rifapentine-containing TB preventive treatment regimen treatment regimens<sup>a</sup> | WHO, 2022 [25] |
| 17 | **Participation in TB vaccine trial** | At least one in-country site is participating in an ongoing or recently completed (last 5 years) Phase IIb/III trial<sup>a</sup> | Treatment Action Group, 2023 [1]<br>Stop TB Partnership, 2023 [2] |

<sup>a</sup> Used an ordinal classification scheme

Responses also led to revisions to the survey language used for several others. Further expert consultation also resulted in the addition of three criteria with objective, publicly available data sources: infant diphtheria-pertussis-tetanus (DPT) coverage as a proxy for immunization capacity and hesitancy, use of 3- or 1-month rifapentine-containing TB preventive therapy regimens as an indicator of early adoption of new TB interventions, and national TB spending to reflect a country’s political will and commitment to TB control. The final list of criteria was also shaped by the availability of objective metrics with country-specific data corresponding to each criterion. Country-specific reference data for each criterion was maintained in an Excel spreadsheet.

### BWS Survey Stakeholder Participants

The main BWS survey to assess the relative importance of decision-making criteria was targeted to stakeholders working in the TB and vaccination fields in countries identified by the project funder, USAID, as priority high TB burden settings. These were: Bangladesh, Cambodia, the Democratic Republic of Congo, Ethiopia, India, Indonesia, Kenya, Kyrgyz Republic, Malawi, Mozambique, Myanmar, Nigeria, Pakistan, Philippines, South Africa, Tajikistan, Tanzania, Uganda, Ukraine, Uzbekistan, Vietnam, Zambia, and Zimbabwe. Stakeholders representing the USAID priority country of Afghanistan were not targeted, given the challenges of conducting research activities in this setting at the current time. The survey also targeted stakeholders working globally and in high TB burden regions (e.g., SSA and Southeast Asia), but not based in the countries named above.

Participants in the sampling frame varied by country but included representatives from community-based or civil society organizations, Ministries of Health, including National TB Program and EPI representatives, national and regional immunization technical advisory groups (NITAGs/RITAGs), professional organizations for health care workers, TB vaccine working groups and networks (e.g., Stop TB Partnership Working Group on New TB Vaccines), research institutions and international nonprofit organizations. The survey was also sent to experts involved in vaccine decision-making and implementation outside of the TB field. Inclusion criteria were: 1) TB vaccine stakeholder as identified by SMART4TB consortium members, USAID or by another external stakeholder; 2) represents a global entity working in TB vaccines or is based in a USAID TB priority country or region; and 3) either represents a TB stakeholder cadre or has expertise in vaccine decision-making and implementation in a field outside of TB. The exclusion criterion was an inability to read/understand English.

To curate the stakeholder list of potential participants, individuals who were on the SMART4TB email distribution list (through affiliation or collaboration with the consortium or by opting to sign up) were organized by high burden country, or regional/global if they worked across country settings. In many cases, SMART4TB members or external TB experts residing in or working closely with one or more of the 23 countries reviewed the list of potential stakeholder participants for a given country. They excluded individuals from the email list – as it includes those working across various TB topic areas or others with a personal or professional interest in TB who would not necessarily be considered a vaccine-specific stakeholder. They also suggested other stakeholders according to the above criteria to ensure a representative mix of potential participants across different stakeholder groups. Additionally, a ‘snowball’ recruitment approach was used whereby survey respondents were asked to provide the names and email addresses for anyone they recommended inviting to complete the survey. If the individual was not already on the distribution list and met the criteria, their email address was added when we sent out a reminder to the larger distribution list, after considering other issues of representation, such as multiple respondents from the same institution or stakeholder group, particularly if the country was close to the maximum target of stakeholders. Furthermore, the USAID global office identified potential respondents working in country USAID missions in target countries.

Given the relatively low response rates anticipated by anonymous electronic surveys, we targeted 5-30 individual stakeholders per country to reach our goal of about 3-5 respondents per country. More populous countries like India and countries with larger TB research programs like Uganda, South Africa, and Zimbabwe were on the high end of the range of targeted stakeholders. Some Central and South Asian countries had fewer identified stakeholders due to smaller TB portfolios, less well-established civil society representation, and English language restrictions.

### BWS Survey Design and Data Collection

The BWS exercise and survey were designed using Sawtooth Software Lighthouse Studio v9.15. The final BWS exercise was a near-perfect balanced incomplete block design where each of the 17 criteria appeared an approximately equal number of times overall (one-way, frequency balance), in conjunction with other criteria (two-way balance, orthogonality), and in each position (position balance). All criteria were shown to participants at least three times to generate reliable individual-level weights. In total, participants were asked to complete 13 randomly different questions (i.e., choice tasks). In each choice task, four criteria were shown at a time, and participants were asked to select what they believed were the most important and least important for determining country selection (see **Figure 2** for an example). To inform their responses, participants were asked to consider a TB vaccine for adults and/or adolescents aligning with current global TB vaccine development efforts [37]. After completing the BWS tasks, participants were asked to directly indicate whether each of the 17 criterion was important for country selection (i.e., a direct anchoring approach) to establish a threshold between important and unimportant criteria [38].

**Figure 2.**
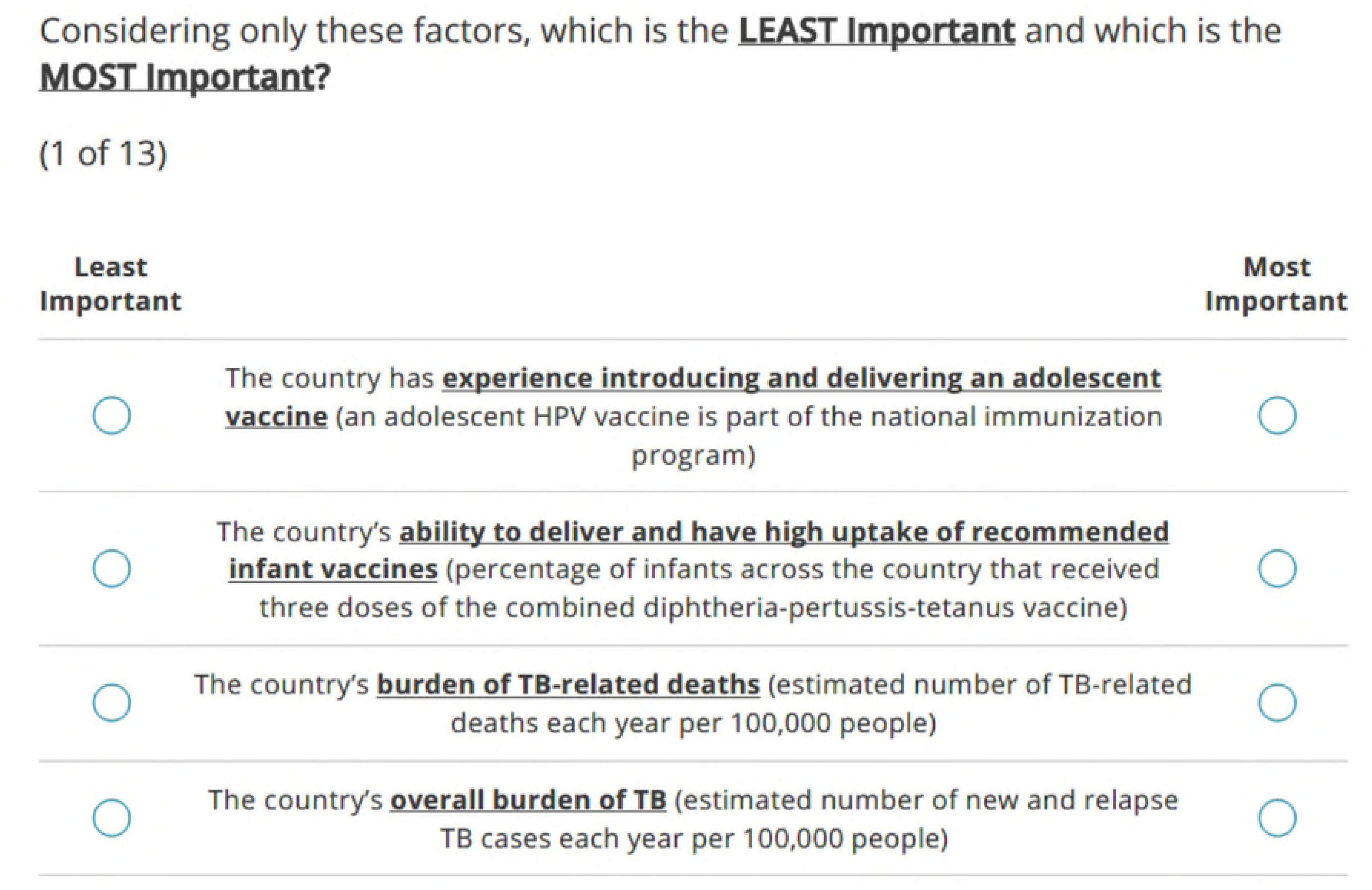
Example of a BWS choice task from the survey. In each choice task, respondents were asked to indicate which one of four criteria they felt was the most important and least important, respectively, for TB vaccine readiness research.

The survey was distributed to potential respondents using MailChimp on July 31, 2023, and the survey closed on August 23, 2023; three emails were sent reminding stakeholders to complete the survey. The email included a brief introduction to the SMART4TB project, a short description of the survey, and a request to complete it. As the study received a non-human subjects research determination and no personally identifiable information was collected, respondents were not required to provide informed consent. However, those clicking on the link in the email were first taken to a page with a longer introduction, which included the purpose of the survey, why they were selected, what they could expect (e.g., duration, type of questions), and who to contact with questions. In addition to the BWS exercise, the survey also captured anonymous background and institution-related information.

### BWS Data Analysis to Estimate Criterion Weighting

The BWS dataset underwent data quality checks, including screening for duplicate responses to ensure each stakeholder’s input was represented only once; none were identified. Next, the Relative Likelihood (RLH) fit statistic was calculated for each participant, which measures how well the predictive model aligns with the observed responses. Sawtooth Software was used to simulate 500 random responders and establish a 95% threshold for the RLH fit statistic specific to the BWS design, a commonly used approach for identifying potentially inattentive or random responders [39]. While 10 participants fell below this threshold, their responses were carefully reviewed for patterns such as extremely fast completion times or contradictory preferences. No such patterns were identified, suggesting valid responses. Therefore, data from all respondents were retained for analysis.

We analyzed the data obtained through direct anchoring in two steps using Hierarchical Bayes (HB) modeling. HB modeling uses an iterative process to estimate parameters, with each iteration refining the estimates based on the data. Compared to other analytic approaches, such as conditional or mixed logit, this approach provides stable individual-level preference weight estimates (i.e., utilities) by assuming each participant’s preferences come from a population distribution while using their specific choices to update both individual and population-level parameters[40]. This was particularly valuable for our diverse stakeholder sample, where preferences might vary systematically across different groups (e.g., policymakers versus advocates) or regions. First, we used an HB model that incorporated this importance threshold to estimate criterion weights relative to the threshold, using 20,000 iterations to ensure stable parameter estimates. Any criterion with a weight (mean of 20,000 iterations) below the threshold was classified as ‘unimportant’ and excluded from further consideration.

We then ran a second HB model without the threshold constraint to calculate criterion weights based only on the BWS choice data, again using 20,000 iterations. From these results, we omitted one criterion identified as unimportant in the first step. The mean weights for the remaining criteria (n=16) were then rescaled at the individual level to sum to 100, making them easier to compare and visualize; these rescaled weights were reported for the BWS-derived estimates. For the final country prioritization score calculations, weights were rescaled again to sum to 1, allowing each country’s final prioritization score to range from 0 to 100 (see below).

### Country-Specific Criterion Scores

Using measurable, predominantly publicly available data (**Table 1**), we scored how each country performed on each criterion using two different approaches depending on the nature of the data. For criteria with continuous data (like TB incidence rates or mortality), we used linear scaling to convert the raw data into comparable scores from 0 to 100. We identified the country with the highest value (assigned 100 points) and the country with the lowest value (assigned 0 points); other countries received scores proportional to where they fell between these endpoints. For criteria representing the presence or absence of specific attributes (e.g., participation in TB vaccine trials, political commitment to end TB), we used a simpler three-point scoring system: 100 points if the attribute was fully present, 50 points if partially present, and 0 points if not present. The final country-specific criterion scores derived from both approaches are shown in **Figure 3**.

**Figure 3.**
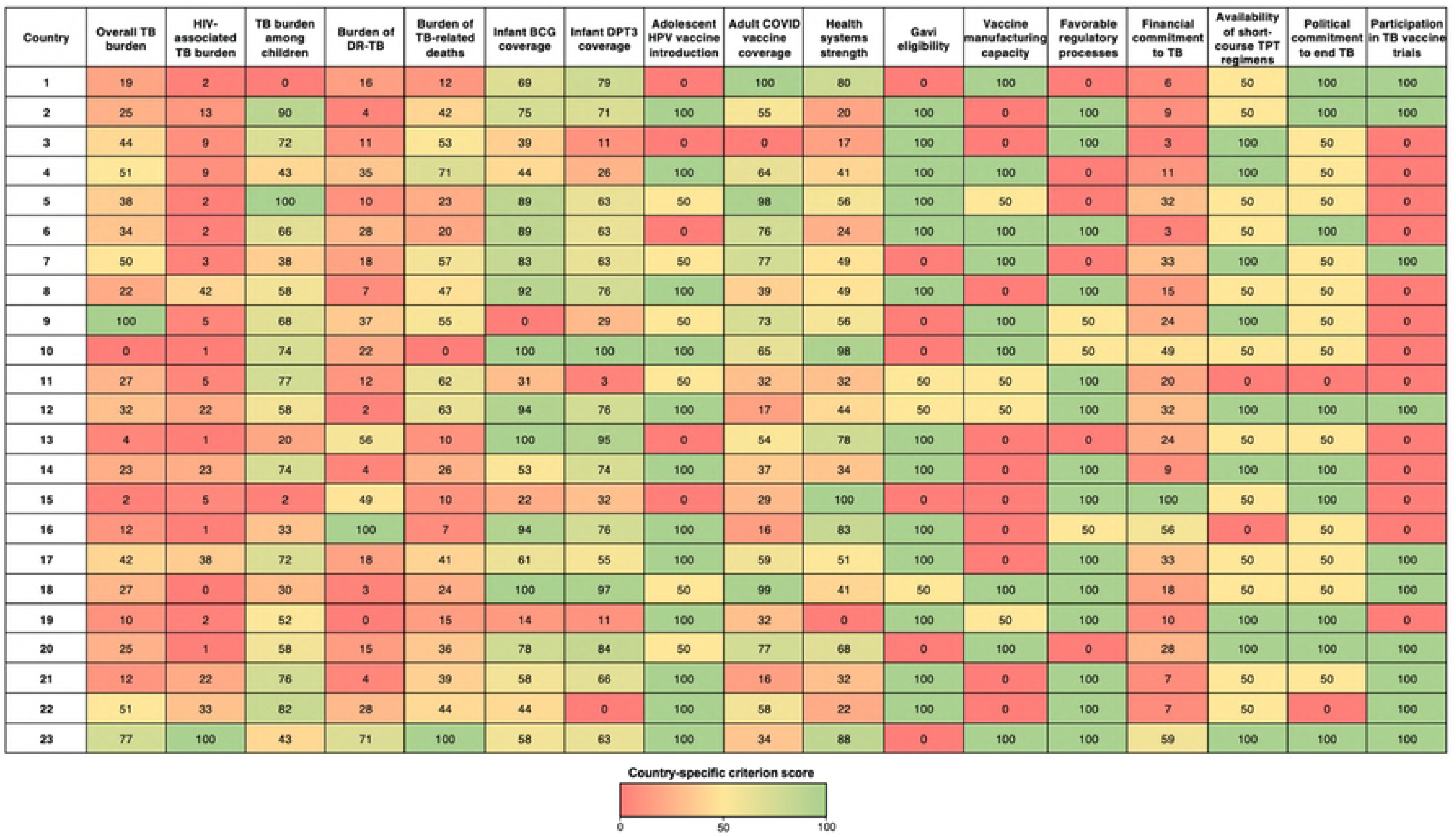
Color-coded matrix of country-specific criterion scores. Color represents the score between 0-100 with lower scores indicated by red and higher scores indicated by green. Note that countries have been anonymized.

### Country Prioritization Score and Country Selection

To determine an overall prioritization score for each country, we first multiplied each country’s criterion score by the corresponding stakeholder-derived weight from the BWS analysis, yielding 16 weighted criterion scores for each country. These 16 scores were then summed to derive the overall prioritization score for each country. While countries were ranked based on these overall scores, we also considered regional diversity to ensure our implementation research would capture insights from different geographic contexts. To balance these objectives while respecting sensitivities around country-specific comparisons, we present our results using anonymized country designations grouped by region.

### Ethics

This study was reviewed by the Johns Hopkins University Institutional Review Board and received a non-human subjects research determination (IRB No. 25465).

## RESULTS

The BWS survey was sent to 427 stakeholders with 115 stakeholders completing the survey (26% response rate). The number of stakeholders targeted and participation rates by country can be found in Supporting Table 1. Among respondents, 101 (88%) were from 23 USAID TB priority countries. There were 17 (74%) countries with ≥ 3 respondents and 22 (96%) countries with ≥ 1 respondent. In the country without any respondents (Myanmar), only one stakeholder was identified to send the survey to versus at least 5 in the other countries.

A diversity of organization types was represented among respondents, with 30% identifying as belonging to a civil society organization or an advocacy group, 13% from National TB Programs, 21% from international non-governmental organizations (NGO), and 17% from academic or research institutions (**Table 2**). With approximately half of the USAID high TB burden countries in SSA, more than half (57%) of the respondents were from this region, while about 17% were from East Asia. Twenty-nine percent of respondents said they were involved in vaccine decision-making in their country, while only 4% of all respondents (including one who indicated being a vaccine decision-maker in their country) were members of NITAGs or RITAGs.

**Table 2.** Survey participant characteristics.

| Characteristic | Total participants (n=115) |
| --- | --- |
| Organization type | Number (Percentage) |
| Civil Society/National NGO | 24 (20.9) |
| International NGO | 24 (20.9) |
| Academic or Research Institution | 20 (17.4) |
| National TB Program | 15 (13.0) |
| Advocacy/Community Organization | 11 (9.6) |
| USAID or Sustaining Technical and Analytic Resources (STAR) Project | 13 (11.3) |
| Other | 8 (7.0) |
| **Region (residence or primarily working in)** |  |
| Sub-Saharan Africa | 66 (57.4) |
| East Asia and Pacific | 19 (16.5) |
| Europe and Central Asia | 15 (13.0) |
| South Asia | 11 (9.6) |
| Other | 4 (3.5) |
| **Key Role** |  |
| Researcher | 25 (21.7) |
| Advocate | 21 (18.3) |
| Program Officer/Coordinator/Administrator/Manager | 19 (16.5) |
| Technical Officer/Adviser/Specialist/Lead | 13 (11.3) |
| Policymaker | 12 (10.4) |
| Clinician | 10 (8.7) |
| Community Organizer | 6 (5.2) |
| Professor | 4 (3.5) |
| Other | 5 (4.3) |
| **Vaccine Decision Maker for Country** |  |
| Yes | 33 (28.7) |
| No | 82 (71.3) |
| **Member of NITAG or RITAG** |  |
| Yes | 5 (4.4) |
| No | 110 (95.6) |

NGO: nongovernmental organization; NITAG: national immunization technical advisory group; RITAG: regional immunization technical advisory group

### BWS Results

Only one of the 17 criteria was identified as ‘unimportant’ for selecting countries for future TB vaccine readiness research - the presence of in-country vaccine manufacturing capacity (see Supporting Table **2**). Following the exclusion of this criterion, the final weights were estimated for each of the remaining 16 criteria (**Figure 4, Supporting Table 3**). A country’s overall TB burden was the most important criterion (weight=11.1), followed by a country’s TB-related political will (weight=10.3). These were significantly more important than the next most important criteria, which were a country’s burden of TB-related deaths (weight=7.9), health system strength (weight=7.5), and adult COVID-19 vaccine coverage (weight=7.4). The least important factors included the burden of HIV-associated TB (weight=3.8), BCG coverage (weight=3.6), and whether a country had participated in a late-stage TB vaccine trial (weight=3.2). A simple count analysis (subtracting the proportion of times each criterion was selected as ‘best’ and ‘worst’ when shown) produced a ranking of criteria that closely aligned with the HB-derived weights (**Supporting Table 4**).

**Figure 4.**
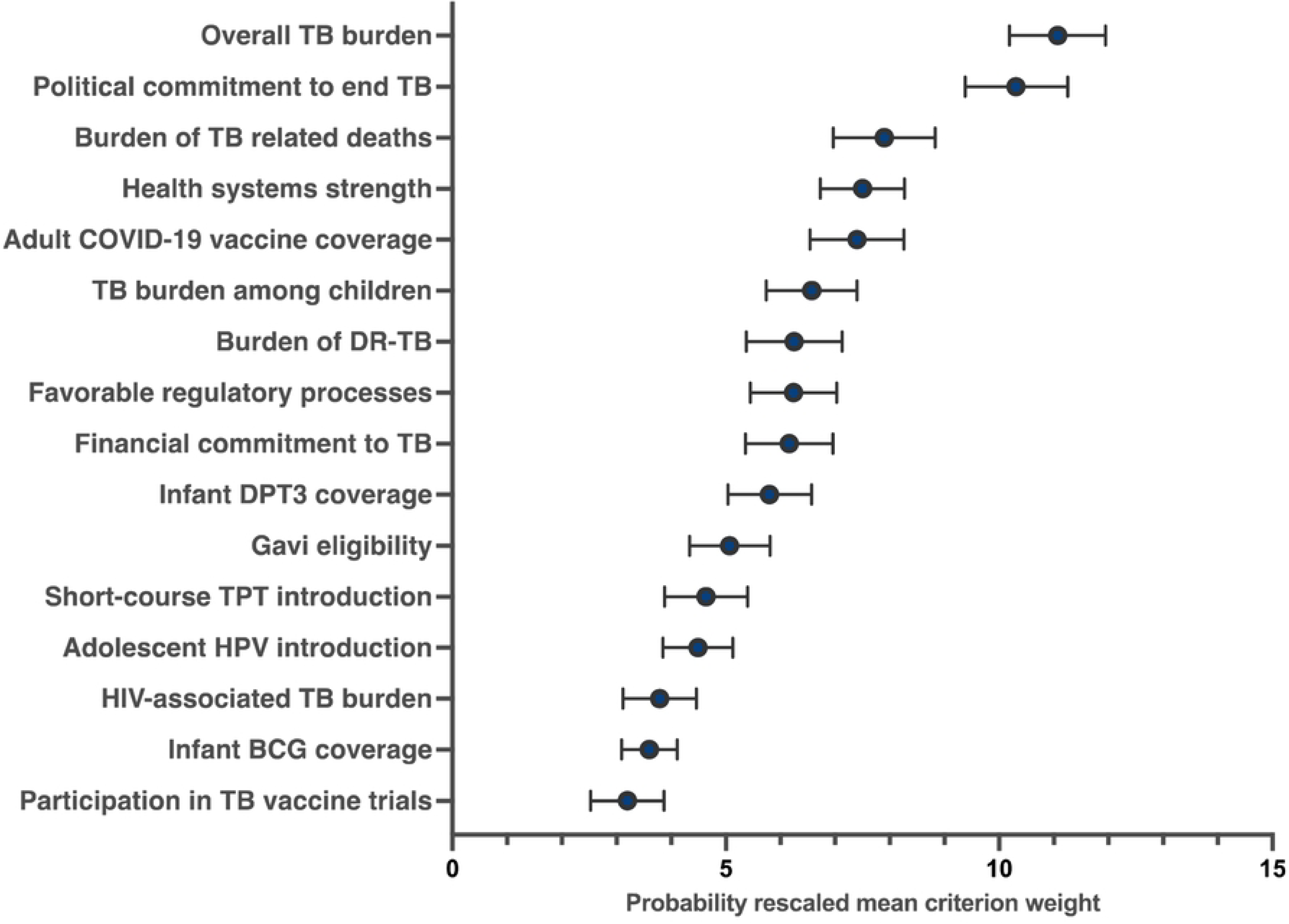
Mean criterion weights and corresponding 95% confidence intervals derived from stakeholder BWS responses to determine the most important criteria for country prioritization for TB vaccine readiness research.

### Country Prioritization Scores and Country Selection

Overall country prioritization scores were derived from the BWS weights and country-specific criteria scores (**Figure 5**). Scores ranged from 32.1 to 74.8, with nine countries exceeding 50 points. While the top five highest-scoring countries were in SSA, our commitment to regional diversification led to the prioritization of the three highest-scoring SSA countries, along with the highest-scoring country from each of these regions: South Asia, Europe and Central Asia, and East Asia and the Pacific.

**Figure 5.**
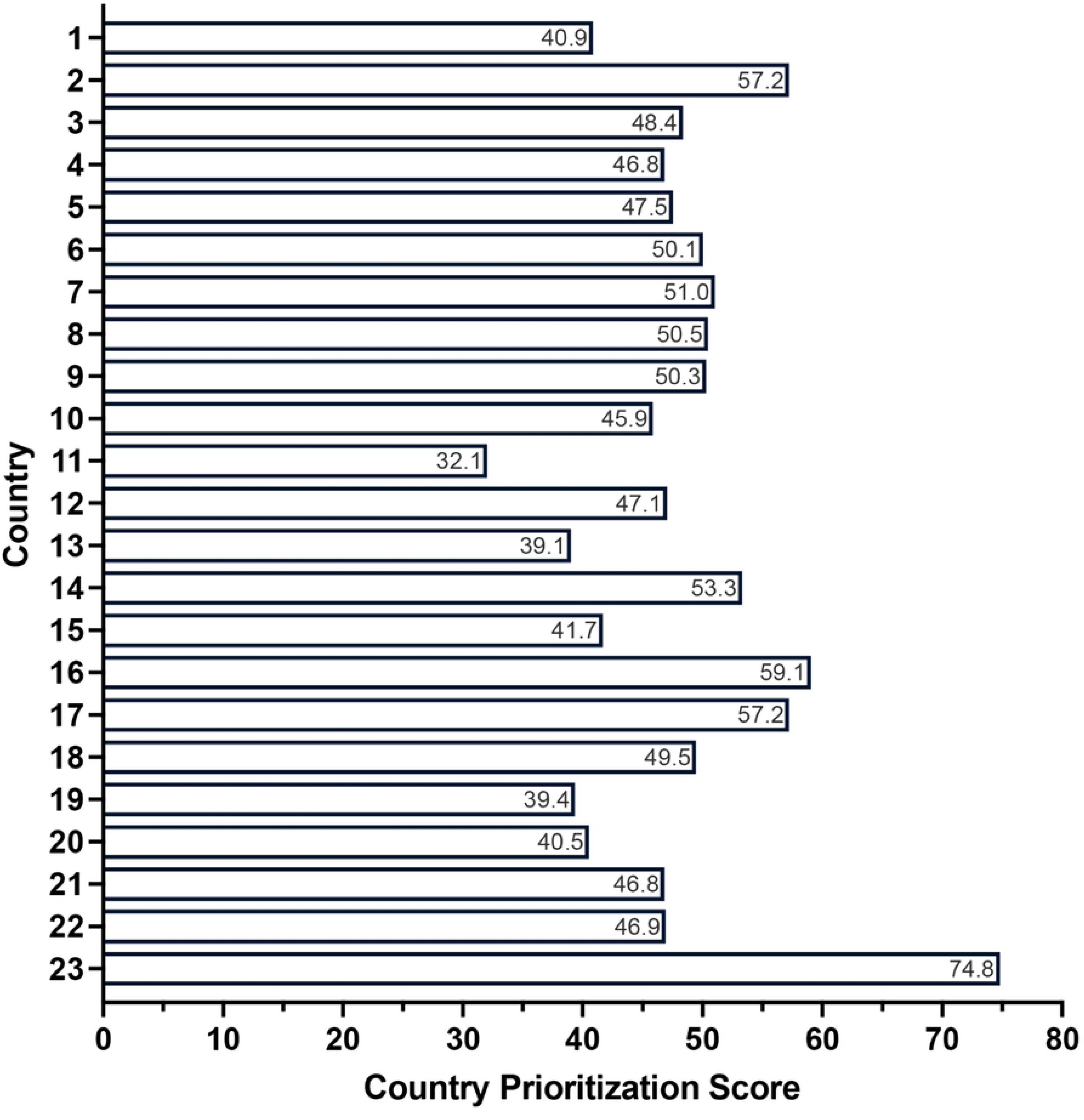
Overall country prioritization scores for selecting countries for future TB vaccine readiness research. Countries have been anonymized.

## DISCUSSION

This study demonstrates how combining stakeholder preferences elicited through BWS with an AHP framework can support country prioritization for TB vaccine readiness research. To our knowledge, this study represents the first application of a hybrid multicriteria decision-making (MCDM) method that combines BWS with the AHP in the public health sector. While other MCDM methods, including standalone AHP, have been utilized in health settings and many other sectors, such as engineering, energy, and environmental sciences, they often present challenges in terms of complexity and intuitiveness [13–15, 41]. This unique integration of BWS and AHP offers a practical, feasible, and transparent approach to effectively address complex decision-making challenges in global health. Our methodology maintains the systematic structure that makes AHP valuable for complex decisions while addressing known limitations. By using BWS to derive criteria weights, we made the process more accessible to diverse stakeholders who might find traditional pairwise comparisons challenging. This modification allowed us to engage stakeholders with varying levels of English-language skills and education, unlikely to have familiarity with decision analysis methods, while still producing robust criterion weights that could be combined with objective country-specific data in a systematic way. It is important to note that while the integration of BWS within the AHP facilitates the inclusion of many individuals in the decision-making process, it does not inherently ensure the engagement of diverse stakeholders. We intentionally sought to include diverse stakeholders, using an approach that was not only relatively easy to implement but also enabled more representative participation and priority setting by individuals with diverse backgrounds, expertise, and perspectives from more than two dozen countries.

While not intended to supplant the complex and often sensitive decision-making surrounding research funding, this stakeholder-engaged process offers an objective process for selection of countries for TB vaccine readiness research and data for consideration alongside other contextual factors. For example, while the AHP-BWS approach identified the highest-scoring six countries as top priorities for research activities, the incorporation of regional diversity meant that the highest-scoring countries were not automatically the final ones selected. This decision reflected our dual goal of facilitating context-specific implementation preparedness for individual countries and enhancing the global relevance of future TB vaccine readiness research. Other contextual factors considered included buy-in from national USAID missions, government willingness to serve as a setting for TB vaccine readiness research, and potential overlap of existing or planned TB vaccine readiness activities (to reduce duplication).

Our findings demonstrate the successful application of this approach, which holds promise for broader use in global public health prioritization. Several key strengths contributed to this successful process. First, we curated stakeholder lists by country - this ensured engagement with a diverse group of participants who possessed differing expertise relevant to TB vaccine implementation, including community advocates, providers, researchers, and policymakers. We assured this through researching backgrounds, engaging known TB experts, and utilizing a snowball approach. Taking into consideration different personal and professional backgrounds allows for a holistic approach to prioritization exercises as individuals rank factors informed by their diverse lived experiences, knowledge, and expertise [8]. Second, we distributed a preliminary survey to a smaller stakeholder group to refine the initial evidence-informed criteria, as well as their corresponding phrasing and data metrics. This step ensured a comprehensive and objectively measured set of criteria for respondents to choose amongst and assess for themselves their relative importance. Finally, utilizing BWS within the AHP, particularly with the added direct anchoring approach, allowed us to determine not only the relative but also the absolute importance of a large number of criteria for each respondent in a user-friendly format [16–18].

A somewhat similar approach using BWS was conducted among multinational TB stakeholders in 14 TB high burden countries (n=33). This study aimed to identify the relative importance of 21 different TB test attributes, seeking to align the development of new diagnostic tests with user and decision-maker needs, and to prioritize preferred features when trade-offs are necessary [42]. Rather than using a simple, direct ranking of importance – which would have been cognitively challenging and potentially inaccurate for so many attributes [19] - respondents were asked several questions in which they selected the most and least important TB test attribute among three randomly generated options. Building on the work of Adepoyibi et al., we sought to address some of the limitations they identified by including a larger sample size. a more diverse set of stakeholder groups, and a broader range of TB high-burden countries. Moreover, we enhanced this approach by integrating the BWS results within the AHP framework, translating BWS-derived weights into actionable insights. This method can help guide decision-making and prioritization among several options; for example, in the case of Adepoyibi et al., determining what specific TB tests should be prioritized for further development based on a prioritization score that combines criterion-specific scores with their respective weights [42]. Existing approaches to prioritization for public health programming and evaluation underscore the need for transparency and stakeholder engagement. While they may consider factors such as disease burden, social aspects, and health impacts, these approaches typically lack a structured framework for explicit prioritization, as noted in a recent review [8]. Our study using AHP integrating BWS offers a promising alternative showcasing its potential applicability of beyond TB vaccine readiness to help address the complexities of global public health prioritization across diverse settings. The methodological framework we’ve developed could be valuable in various public health contexts requiring structured prioritization with diverse stakeholder input. This approach could help decision-makers prioritize among competing options at any level - from selecting intervention sites within a single country to choosing between different service delivery models or allocating limited resources across multiple programs. The framework is especially suited to scenarios where decisions must balance multiple factors while ensuring meaningful input from varied stakeholders. Building on this global stakeholder consensus process, we will now undertake implementation research in the six prioritized high TB burden countries to better understand factors influencing TB vaccine acceptance and uptake among TB-affected communities and the readiness of health systems to implement and sustain equitable delivery of future TB vaccines.

While our study advances the methodology of public health prioritization and has practical implications for subsequent research in TB vaccine introduction and implementation, it has limitations. The survey link, accessible to anyone if forwarded, could have extended beyond our target audience; despite this, we made concerted efforts to achieve a representative distribution list in terms of both country and stakeholder type. Also, since we did not collect personal information, directly matching respondents to the distribution list was not possible. Nonetheless, we performed multiple checks to reduce the likelihood of duplicate responses. Further, the respondent demographics were fairly well distributed among different stakeholder groups, mitigating potential biases in the outcomes due to representation imbalances. However, any imbalances in representation (e.g., by country) would not bias the results in a particular direction or towards a certain country, given the indirect preference elicitation nature of BWS, coupled with the fact that country-specific criterion scores were unknown to respondents. Additionally, the survey’s reach was limited to English-comprehending individuals with internet access.

Snowball sampling could have exacerbated this bias and led to greater representation of certain stakeholder groups (e.g., NGOs or academic institutions), though this was considered and not every recommended stakeholder was included in a subsequent survey invitation. This limitation was also balanced with the opportunity it provided for broad reach to people in over two dozen predominantly high TB-burden countries by bypassing the need for interviewer-facilitated survey administration. Due to a truncated timeline, we were unable to conduct pilot testing of the BWS and its 13 choice tasks that would have been valuable to assess potential respondent fatigue and comprehension issues prior to full implementation. However, we chose BWS over traditional pairwise comparisons in AHP to improve accessibility and reduce cognitive burden, and quality metrics (e.g., RLH-values) suggest the impact was likely minimal. Furthermore, the final country rankings were kept anonymous. We are mindful that the objective and transparent nature of our method for country prioritization may still raise potential concerns and sensitivities due to the comparative assessment of countries, particularly those that could be interpreted as reflecting the quality of healthcare delivery. Thus, the anonymization of results was not intended to obscure the findings, and ultimately, we do not feel that it undermines the lessons that can be learned from our experience.

In conclusion, our study demonstrates the feasibility and value of an inclusive and objective approach to public health prioritization, in this case, in the context of TB vaccine readiness research. This work offers a replicable framework for addressing similar challenges across the spectrum of global health initiatives. Given the complexities of ongoing and emerging global health challenges, there is significant value in adopting this approach to planning, implementation, and evaluation.

## Data Availability

All data generated or analyzed during this study are included in the submitted manuscript and its supporting information files (dataset to be provided upon acceptance of the paper).

## Acknowledgements

The authors would like to thank all stakeholders who completed the survey and other SMART4TB collaborators who contributed to the success of this survey. We would also like to acknowledge Rekha Radhakrishnan and Isadora Salles for their support with stakeholder communication and data management, respectively.

## Supporting Information

1. **S1 Table.** Stakeholder recruitment by country
2. **S2 Table.** Mean criterion weights and corresponding 95% confidence intervals for the 17 prioritization criteria derived from stakeholder BWS responses and incorporating a direct anchoring approach
3. **S3 Table.** Mean criterion weights and corresponding 95% confidence intervals for the 16 final prioritization criteria deemed important derived from stakeholder BWS responses.
4. **S4 Table.** Best Worst Scaling Count Analysis (n=115)

